# Health impacts of COVID-19 disruptions to primary cervical screening by time since last screen: A model-based analysis for current and future disruptions

**DOI:** 10.1101/2022.07.25.22278011

**Authors:** Emily A. Burger, Inge M.C.M. de Kok, James F O’Mahony, Matejka Rebolj, Erik E.L. Jansen, Daniel D. de Bondt, James Killen, Sharon J. Hanley, Alejandra Castanon, Jane J. Kim, Karen Canfell, Megan A. Smith, Mary Caroline Regan

## Abstract

**Background:** We evaluated how temporary disruptions to primary cervical cancer (CC) screening services may differentially impact women due to heterogeneity in their screening history and test modality.

**Methods:** We used three CC models to project the short- and long-term health impacts assuming an underlying primary screening frequency (i.e., 1, 3, 5, or 10 yearly) under three alternative COVID-19-related screening disruption scenarios (i.e., 1-, 2- or 5-year delay) versus no delay, in the context of both cytology-based and HPV-based screening.

**Results:** Models projected a relative increase in symptomatically-detected cancer cases during a 1-year delay period that was 38% higher (Policy1-Cervix), 80% higher (Harvard) and 170% higher (MISCAN-Cervix) for under-screened women whose last cytology screen was 5 years prior to the disruption period compared with guidelines-compliant women (i.e., last screen three years prior to disruption). Over a woman’s lifetime, temporary COVID-19-related delays had less impact on lifetime risk of developing CC than screening frequency and test modality; however, CC risks increased disproportionately the longer time had elapsed since a woman’s last screen at the time of the disruption. Excess risks for a given delay period were generally lower for HPV-based screeners than for cytology-based screeners

**Conclusions:** Our independent models predicted that the main drivers of CC risk were screening frequency and screening modality, and the overall impact of disruptions from the pandemic on CC outcomes may be small. However, screening disruptions disproportionately affect under-screened women, underpinning the importance of reaching such women as a critical area of focus, regardless of temporary disruptions.

**Funding:** This study was supported by funding from the National Cancer Institute (U01CA199334). The contents are solely the responsibility of the authors and do not necessarily represent the official views of the National Cancer Institute. Megan A Smith receives salary support from the National Health and Medical Research Council, Australia (APP1159491) and Cancer Institute NSW (ECF181561). Matejka Rebolj is funded by Cancer Research UK (reference: C8162/A27047). James O’Mahony is funded by Ireland’s Health Research Board (EIA2017054). Karen Canfell receives salary support from the National Health and Medical Research Council, Australia (APP1194679). Emily A. Burger receives salary support from the Norwegian Cancer Society.

## INTRODUCTION

The coronavirus disease 2019 (COVID-19) pandemic continues to impact on a wide range of health outcomes. In the initial months of the pandemic in 2020, there were severe disruptions to preventive services including cervical cancer screening. For example, during acute phases of the pandemic in the United States (U.S.), 59% of federally qualified health centers stopped cancer screenings completely (1), and electronic health records from 39 organizations spanning 23 States found a 67% decline in mean weekly cervical cancer screening volumes (2). While cancer screening volumes gradually improved (2), mid-June 2020 volumes remained around 30% lower than their pre-COVID-19 levels, and cervical volumes have remained 10% lower two years into the pandemic (3).

The risk of developing cervical cancer depends in part on time since a last screen (4, 5). Despite U.S. recommendations for primary cervical cancer screening of either 3-yearly cytology or 5-yearly HPV testing (6), there is heterogeneity in adherence to guideline-recommendations where both under-screening and over-screening are observed, when comparing behavior to recommendations. For example, in the only population-based registry in the U.S. prior to widespread primary HPV-based screening, 20% of women were not screened within 5 years (7), which was correlated with race and ethnicity, income level, lower levels of education and lack of insurance (8). Conversely, screening more frequently than recommended has been observed in 66% of insured women (9).

The impact of service disruptions due to COVID-19 may not have affected all women equally. For women without health insurance or unable to access care, or those who avoid care due to fear of COVID-19, the disruptions may continue. In other countries such as the United Kingdom (U.K.), 30% of survey respondents elicited Fall 2020 reported that they were less likely to attend cervical screening now than before the pandemic (10). Although the observed decrease in screening attendance ultimately was smaller than surveyed intentions to screen (11), the U.K. study also found that previous non-participation was the strongest predictor of low intentions for future post-pandemic participation.

Rebounds towards pre-pandemic attendance levels in aggregate-level metrics may suggest a successful recovery but could actually mask unexpected disparities in coverage. For example, the same disruption period may differentially impact women due to heterogeneity in their screening history so that the impact is greater for those under-screened compared to those that are screened according to recommended guidelines. It will thus be important to understand the influence of variation in women’s past behavior as a contributor to underlying risk when assessing past and ongoing disruptions to screening.

Empirically, decreases in cervical cancer diagnoses in 2020 have been confirmed in the U.S. (12) and elsewhere (13). Previous model-based analyses have projected that temporary disruptions to cervical cancer screening may result in temporal shifts in cancer detection (initial decreases followed by an increase), yielding small net increases in cervical cancer burden (14, 15). Such decreases are to be expected in the short run due to the reduction in screening and related investigations and any net increases will only be observed in time. Model-based analyses (14, 15) have shown that maintaining services for the highest risk women may mitigate the potential secondary impacts of COVID-19 on cervical cancer; for example, prioritizing those in need of surveillance, colposcopies or excisional treatment, as well as women whose last primary screen did not involve a highly sensitive test, such as that for the detection of human papillomavirus (HPV). Furthermore, short delays to cervical screening services among women with a previous negative HPV result had minor effects on cancer outcomes; however, previous analyses have not explicitly stratified outcomes for women by their prior screening history, i.e., time since last screen.

Disease simulation models can help assess the impact of service disruptions and policy responses in advance of empirical data. Models can also quantify health consequences of alternative screening disruption scenarios and isolate complex interactions between temporary screening suspensions for women with different underlying screening histories. These simulations can help inform which women are most vulnerable to COVID-19 disruptions and should be prioritized for targeted recovery activities. Therefore, as part of the Covid and Cancer Global Modelling Consortium (CCGMC), we used three US-contextualized cervical cancer natural history models from the Cancer Intervention and Surveillance Modeling Network (CISNET) consortium (https://cisnet.cancer.gov/) to isolate the health impact of temporary disruptions to *primary screening services only* by time since a woman’s last screen and primary screening test modality. Multi-model comparative analyses can demonstrate the validity of findings and test the robustness despite structural differences between the models used. The purpose of this analysis is to provide decision makers with evidence regarding the potential impact of temporary disruptions to the provision of screening services on cervical cancer incidence, either due to the COVID-19 pandemic or any other similar disruption, on a disaggregated basis according to women’s prior screening history in order to inform any targeted allocation of scarce screening capacity.

## METHODS

### Analytic overview

To complement our previous analysis (14), we used the same three CISNET-Cervical microsimulation models to project the expected lifetime risk (until age 84 years) of developing cervical cancer for three birth cohorts (born in 1965, 1975 and 1985; aged 55, 45 and 35 in 2020, respectively) assuming an underlying exposure to HPV vaccination (see (16)) and screening frequency—that is, annual, 3-yearly, 5-yearly, or 10-yearly screening, aligned so that 2020 was 1, 3, 5, or 10 years since their last screen (**Figure 1; Appendix Tables A1 and A2**). These selected birth cohorts enabled the analysis to capture at least 10 years of pre- and post-COVID-19 screening history. We used the models to estimate both the short- and long-term impacts of COVID-19 delays on cervical cancer burden. As both primary cytology- and HPV-based screening modalities are recommended in the U.S., we explored these outcomes in the context of primary cytology (i.e., Pap smear only) with and without switching to primary high-risk HPV testing from age 30 years with partial genotyping for HPV genotypes 16 and 18. Screen-positive women were managed according to guidelines (17) and followed Kaiser Permanente Northern California compliance patterns, i.e., colposcopy compliance (79%), precancer treatment compliance (73%) (18). Scenarios were simulated in the context of birth cohort-specific historical HPV vaccination coverage as estimated and applied in another analysis (19).

**Figure 1.**
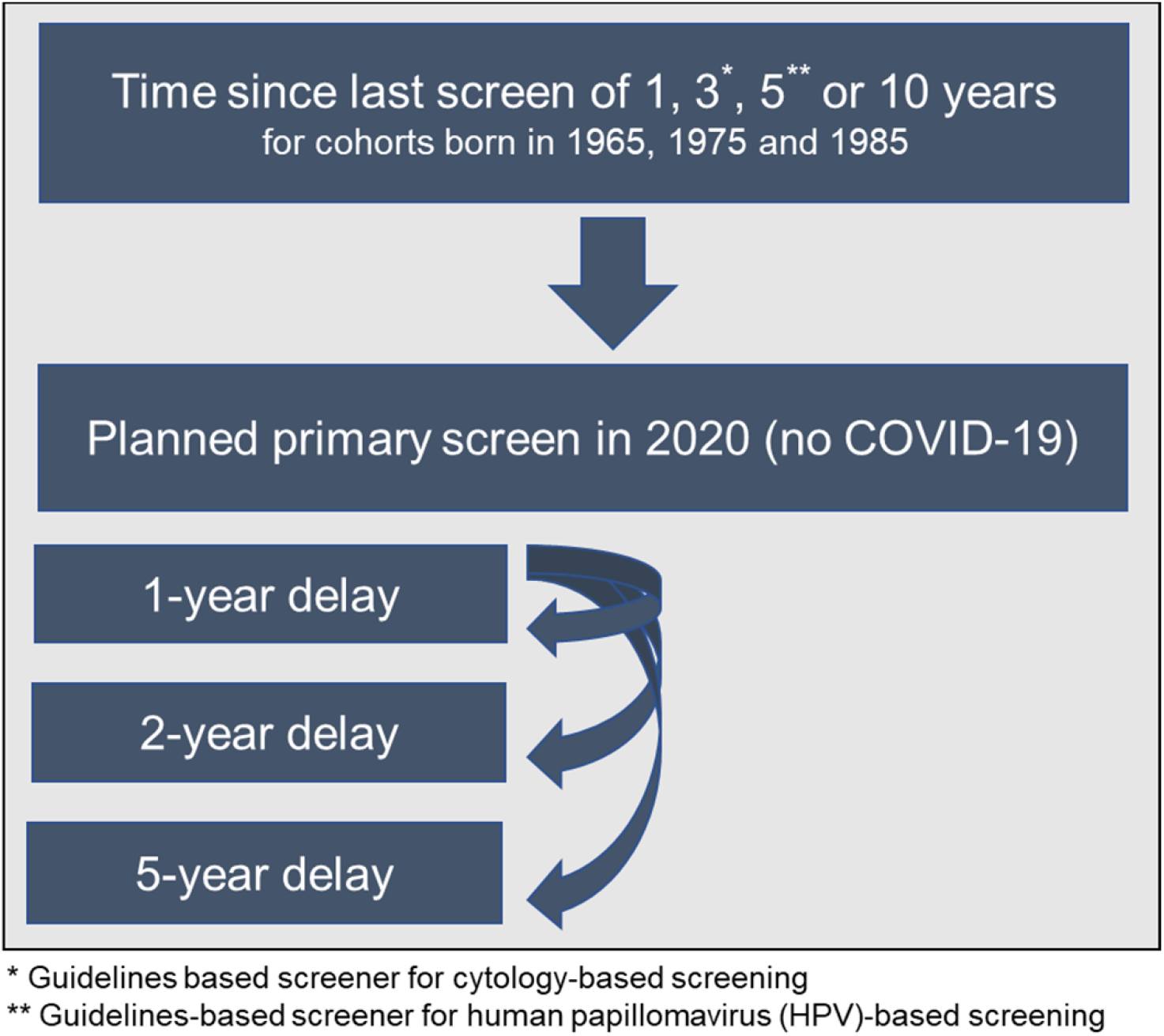
Scenario overview reflecting the heterogeneity in screening history (aligned so that 2020 was 1, 3, 5, or 10 years since their last screen) facing alternative COVID-19 delay disruptions for three birth cohorts of women.

For the short-term impacts, we estimated the relative excess rate of symptomatically-detected cancer during a COVID-19-related screening delay period for under-screeners compared with “guidelines-compliant screeners” (i.e., those who perfectly adhere to 3-year cytology screening or 5-yearly HPV screening, except for during the Covid-19 disruption period). We averaged the cancer incidence rates (per 100,000 women) accumulated during a delay period (i.e., 1, 2 or 5 years) across the three birth cohorts for each screening history profile (i.e., 3-yearly, 5-yearly or 10-yearly screening). The denominator for each relative rate (RR) calculation was the accumulated cancer rate under a given delay period for a guidelines-compliant screener, which differed according to primary test modality (i.e., 3-yearly screening for cytology-based screeners and 5-yearly screening for HPV-based screeners) (**Appendix Figure A1**). For the long-term impact, we projected the impact of disruptions to lifetime risks and absolute changes in cancer risks for each of the three alternative COVID-19-related screening delay scenarios, compared to a scenario of no COVID-19-related disruptions. To set findings within the wider context of prevention, we additionally considered how much each scenario would reduce a woman’s lifetime risk of developing cervical cancer (compared to a hypothetical no screening scenario). For each scenario, model projections of cervical cancer cases and lifetime risks were averaged across the three birth cohorts.

### Simulation models

As previously described (14, 20, 21), the three CISNET-Cervical models (Harvard, MISCAN-Cervix and Policy1-Cervix) reflect the natural history of HPV-induced cervical cancer but differ structurally with respect to the type and number of health states, HPV genotype categorizations, histological cancer types, model cycle length and data sources used to parameterize the model prior to fitting to the U.S. population. Standardized US-model inputs included hysterectomy rates, all-cause mortality, and cervical cancer survival (20). To reflect the burden of HPV and cervical cancer in the U.S., the models were calibrated to HPV and cervical disease outcomes, achieving good fit to empirical targets based on U.S. women (See Burger et al 2020 (20) for details of the calibration and fitting processes).

### Scenarios and assumptions

We assumed that in the absence of the COVID-19 pandemic, each cohort would have received a primary cervical screen in 2020, aligned with an underlying screening frequency, i.e., 1, 3, 5, or 10 years since the last screen. For each birth cohort and screening frequency combination, these women faced either no delay, or a 1-, 2- or 5-year delay (**Figure 1, Appendix Table A2**). We assumed that during the delay period, there was a 100% temporary loss in primary screening, but following the delay period, screening was assumed to immediately resume, and women would continue to follow their pre-pandemic screening frequency. We assumed COVID-19 did not impact attendance for surveillance, diagnosis, or treatment of screen-detected abnormalities or investigation for symptomatically-detected cancers, except when directly implied by missed screening events during a COVID-19 delay period. In line with U.S. guideline recommendations, all models assumed women did not attend for routine screening after age 65 years. A key modifier of the impact of screening delays on lifetime risk is the age at which women received their last screening test (22). Due to the analytically fixed screening intervals assumed post-COVID-19-related delay, the timing of future screening was shifted in all cohorts other than annual screeners; as a result, for some combinations of screening frequency and COVID-19-related delays, the delays also reduced the number of lifetime screens and/or changed the age at last screen (**See Appendix Tables A1 and A2 for additional details**). For example, for a woman born in 1975 who screens every 10 years, her last screen would be at age 65 years without a COVID-19 disruption; however, her last screen would occur at age 56, 57 or 60 under the 1-, 2- or 5-year delay scenarios.

## RESULTS

### Short-term impacts

On average, among women aged 35-55 years, the models projected a relative increase in symptomatically-detected cancer burden during a 1-year delay period that were higher – 38% higher (Policy1-Cervix), 80% higher (Harvard) and 170% higher (MISCAN-Cervix) – for those who had not screened in 5 years at the time of the disruption, compared with women who attended cytology-based screening according to guidelines (i.e., every three years) (**Figure 2; left panels**). Compared with guidelines-compliant cytology screeners, the relative excess burden of cancers detected during a 1-year delay period was 3.1 (Policy1-Cervix), 3.2 (Harvard), or 7.0 (MISCAN-Cervix) times higher for women whose last cytology screen was 10 years ago at the time of the disruption. Compared with women who switched to HPV-based screening at age 30 and were guidelines-compliant screening every 5 years, women who screened every 10 years with HPV after age 30 years faced an excess cancer burden that was generally consistent regardless of the disruption period, ranging from 2.2-2.5 (Policy1-Cervix), 3.0-3.9 (MISCAN-Cervix), and 3.6-3.7 (Harvard) times higher (**Figure 2; right panels**). Although the relative excess burden among women overdue for screening remained generally similar by delay period, the absolute accumulated rates increased with the length of the delay period (**Appendix Table A3**).

**Figure 2.**
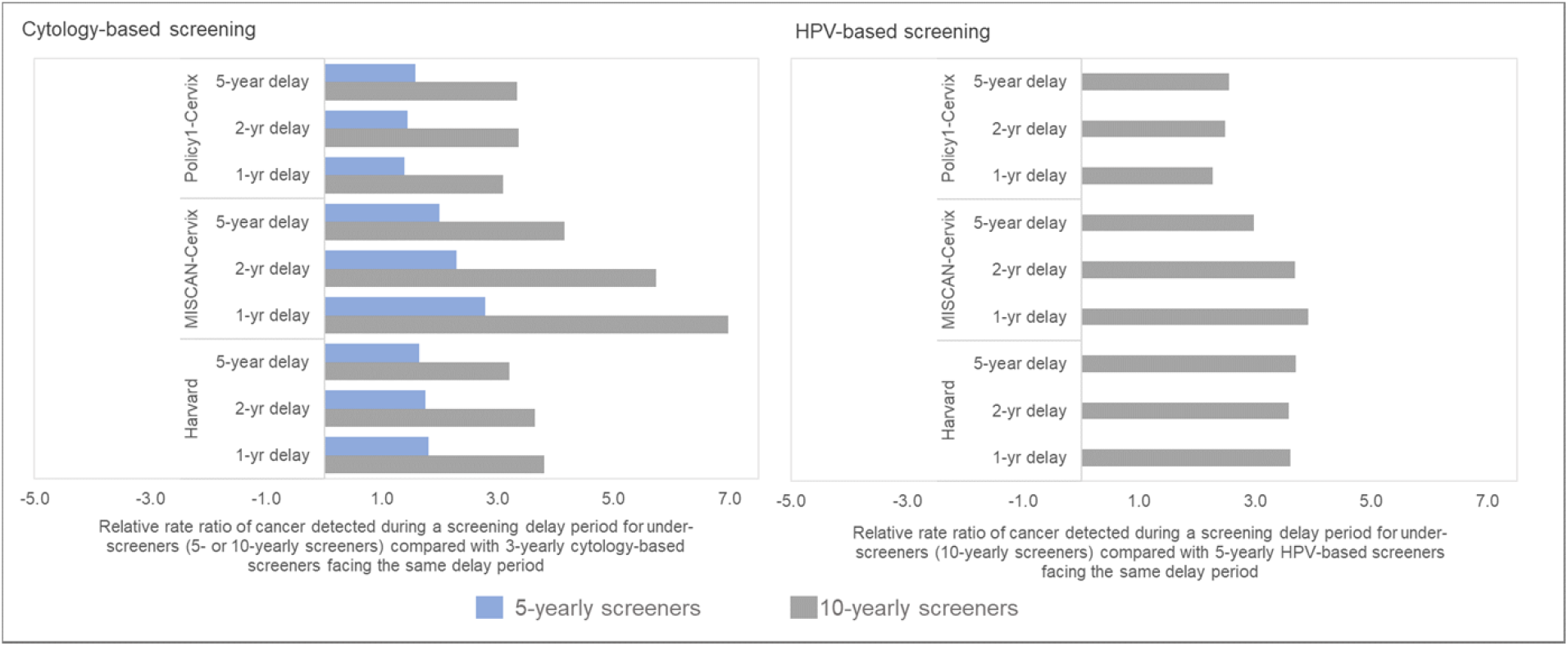
Short-term impacts: Relative rate ratio of cancer detected during the screening delay period for under-screeners compared with the same delay duration for guidelines-compliant screeners.

### Long-term impacts

The models consistently projected that a 1- or even 5-year temporary disruption to primary screening had a smaller effect on the lifetime risk of developing cervical cancer than the effects from screening frequency and modality we considered (**Figure 3**). For example, within-model comparisons found that the lifetime risk of developing cervical cancer was lower, even in the context of an extreme 5-year screening disruption, for women screened every 3 years with cytology prior to the disruption (Policy1-Cervix (0.21%) Harvard (0.32%), and MISCAN-Cervix (0.32%)) than for women screening every 5 years without a disruption (Policy1-Cervix (0.22%), Harvard (0.42%), and MISCAN-Cervix (0.39%)) (**Figure 3, upper panels**). Set within the wider context of prevention, the models projected that, under an extreme scenario of a 5-year delay, 3-yearly cytology screening maintained nearly all benefits of screening, decreasing from preventing 72.1% to 69.1% (MISCAN-Cervix), 79.9% to 78.4% (Harvard), and 86.5% to 85.1% (Policy1-Cervix) of cancer cases over a woman’s lifetime compared with no screening (assuming screening resumed following the disruption) (**Appendix Table A4)**. In contrast to women screened with cytology over their lifetime, women screened with primary HPV after age 30 years face a lower overall lifetime risk of cervical cancer (and percentage of cancers prevented by screening was higher) compared with cytology-based screening; furthermore, these women generally faced smaller impacts of a COVID-19 disruption to screening relative to screening frequency (**Figure 3, right panel; Appendix Figure A4**).

**Figure 3.**
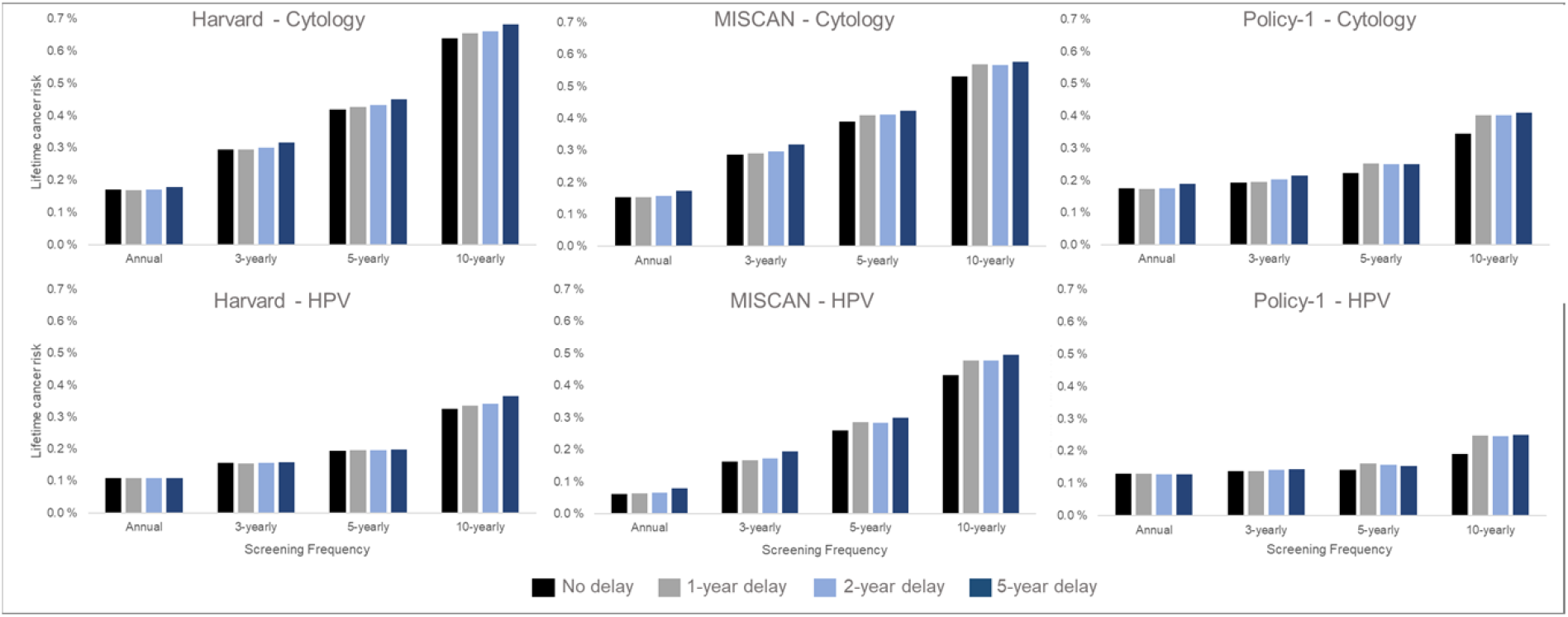
Long-term impacts: Projected impact of COVID-19-related disruptions to primary cervical cancer screening on the lifetime risk of developing cervical cancer (averaged across the 1965/1975/1985 birth cohorts of women) by time since last screen for cytology-based screening (top panels) and human papillomavirus (HPV)-based screening (bottom panels) for three CISNET-Cervical disease simulation models.

Despite the relatively lower contribution of COVID-19-related delays to lifetime risk of developing cervical cancer than screening frequency and test modality, there were important differences in the impact of a delay period on a woman’s lifetime risk by the length of time since her last screen and the screening modality used during the last screen (**Figure 4**). In general, annual or 3-yearly screeners faced only nominal excess risks when experiencing a 1-year temporary delay to primary cytology screening, and cancer risks increased disproportionately the longer time had elapsed since a woman’s last screen (**Figure 4; upper panels**). For example, all models projected that compared with no COVID-19 delay, an extreme 5-year temporary delay scenario was expected to increase the number of remaining lifetime cervical cancer cases by 20 (Policy1-Cervix), 22 (Harvard), and 31 (MISCAN-Cervix) per 100,000 women screened 3-yearly with cytology, compared to an increase of 44 (Harvard), 47 (MISCAN-Cervix), and 66 (Policy1-Cervix) per 100,000 women screened 10-yearly with cytology (**Table 1**). Importantly, these excess risks for a given delay period were generally lower for HPV-based screeners than for cytology-based screeners (**Figure 4; lower panels**). For example, compared with cytology-based screening, two of the models (Harvard and Policy1-Cervix) found that woman screened with primary HPV testing faced smaller excess risks for the same delay duration unless women were screening very infrequently (10-yearly), in which case, the excess risks of cancer were similar, i.e., 44-66 per 100,000 women for cytology versus 40-58 per 100,000 women for HPV 100,000 women).

**Table 1.** Long-term health impacts* of a 5-year temporary delay to screening compared with no delay, by screening history, i.e., screening frequency.

|  |  | Screening frequency |  |  |  |
| --- | --- | --- | --- | --- | --- |
|  |  | Annual | 3-yearly | 5-yearly | 10-yearly** |
| **Primary cytology-based screening** |  |  |  |  |  |
| *Harvard* |  |  |  |  |  |
| Absolute change in lifetime risk |  | 0.008% | 0.022% | 0.031% | 0.044% |
| Excess cases over lifetime per 100,000 women |  | 8 | 22 | 31 | 44 |
| *MISCAN-Cervix* |  |  |  |  |  |
| Absolute change in lifetime risk |  | 0.021% | 0.031% | 0.034% | 0.047% |
| Excess cases over lifetime per 100,000 women |  | 21 | 31 | 34 | 47 |
| *Policy1-Cervix* |  |  |  |  |  |
| Absolute change in lifetime risk |  | 0.014% | 0.020% | 0.029% | 0.066% |
| Excess cases over lifetime per 100,000 women |  | 14 | 20 | 28 | 66 |
| **Primary HPV-based screening** |  |  |  |  |  |
| *Harvard* |  |  |  |  |  |
| Absolute change in lifetime risk |  | 0.000% | 0.001 % | 0.004 % | 0.040% |
| Excess cases over lifetime per 100,000 women |  | 0 | 1 | 4 | 40 |
| *MISCAN-Cervix* |  |  |  |  |  |
| Absolute change in lifetime risk |  | 0.017 % | 0.032% | 0.041% | 0.063% |
| Excess cases over lifetime per 100,000 women |  | 17 | 32 | 41 | 63 |
| *Policy1-Cervix* |  |  |  |  |  |
| Absolute change in lifetime risk |  | 0.000% | 0.006% | 0.012% | 0.058% |
| Excess cases over lifetime per 100,000 women |  | 0 | 6 | 12 | 58 |

\*Risks are rounded to nearest 0.001%; \*\*The women born in 1985 (aged 35 in 2020) received their last screen at age 25 and have not yet made the switch to primary human papillomavirus (HPV)-based screening. In the
primary HPV-based analysis, these women would switch to primary HPV-based screening for their remaining lifetime either at 35 (under the no delay scenario) or aged >35 years (with a delay).

**Figure 4.**
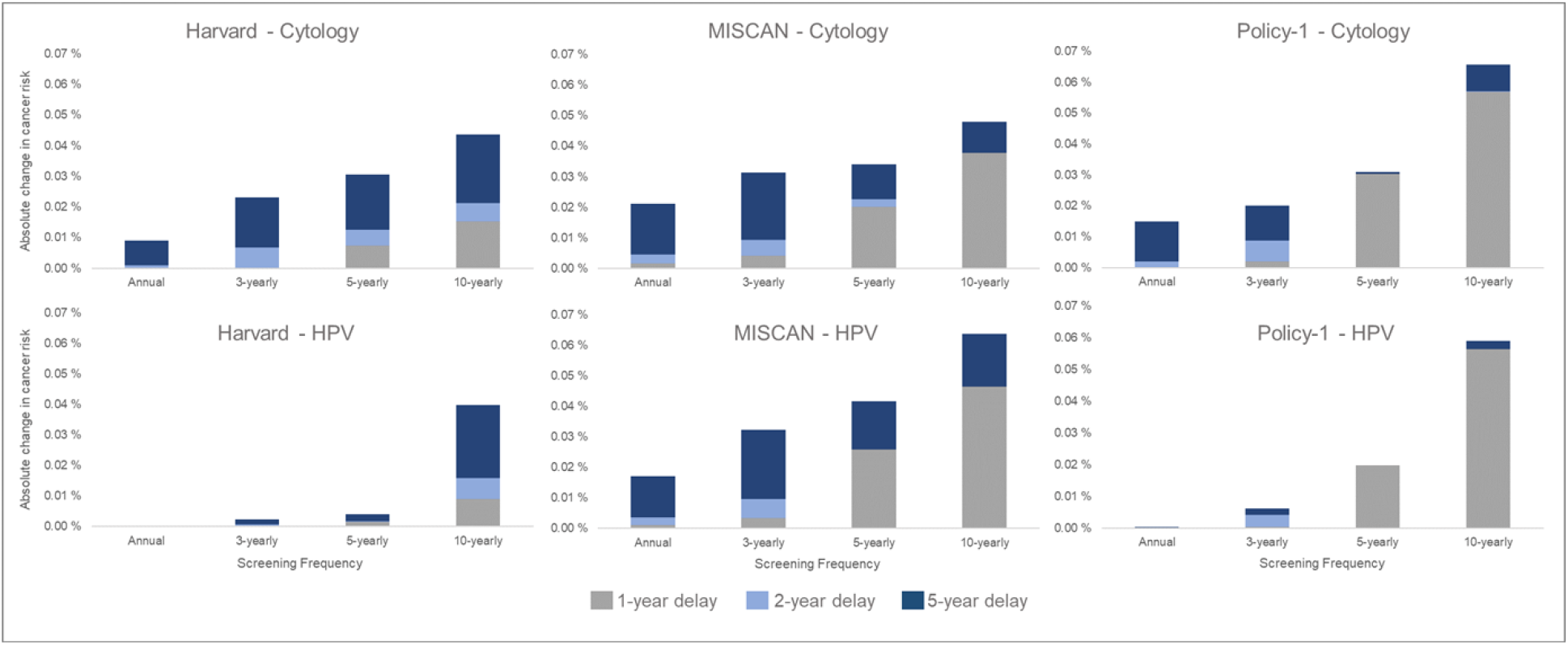
Long-term impacts: Projected impact of COVID-19-related disruptions to primary cervical cancer screening on the incremental lifetime risk of developing cervical cancer (averaged across the 1965/1975/1985 birth cohorts of women) by time since last screen for cytology-based screening (top panels) and human papillomavirus (HPV)-based screening (bottom panels) for three CISNET-Cervical disease simulation models.

## DISCUSSION

The results of our comparative health impact modeling study suggest women who are overdue for screening and encounter a further delay face an increased risk of presenting with a symptomatic cancer during the delay period compared with those who attend screening regularly according to guidelines. Furthermore, women who undergo routine guidelines-compliant screening are able to endure a temporary, i.e., 1-year, disruption to cervical screening under both primary cytology and HPV-based screening modalities, and those who undergo guidelines-compliant screening with HPV testing are more resilient to longer delays (2 or 5 years). In the U.S., where there is heterogeneity in screening behavior (9), our findings suggest that to minimize population cancer risk, targeted outreach to over-screened and regularly-screened women should be a lower priority than outreach to women whose screening history is not up-to-date. Our findings support outreach to women most vulnerable to COVID-19 disruptions who also faced pre-pandemic barriers to routine screening. Importantly, aggregated metrics demonstrating a near-return to pre-pandemic screening volumes may not be adequate to capture heterogeneities in screening history, and therefore, risk associated with disruptions to screening.

Similar to our previous results (14), we found that screening with primary HPV testing generally provided greater reductions to lifetime risk of developing cervical cancer compared with cytology-based screening. Although we did not explicitly simulate screening that switches from cytology-based screening before a disruption to HPV testing upon resumption such a strategy may be able to mitigate any pandemic-related excess risks. Similarly, we found that the impact of disrupting an HPV-based screening program has different implications than the disruption of a cytology-based program. This can be explained by the fact that HPV screening has a higher sensitivity to detect (pre-invasive) cervical lesions; therefore, the cancer risk at time of disruption is lower and this may provide a greater buffer to endure temporary disruptions. On the other hand, in case of the more sensitive HPV test, disruption takes away a relatively more valuable screening moment. The balance between these two factors causes a greater or smaller excess risk per delay duration in case of HPV screening compared to cytology screening. If in a model the first effect is larger than the second effect, disruption of the HPV program has a smaller effect than of the cytology program, which is the case for all screening frequencies in both the Harvard and Policy1-cervix models and the annual screeners in MISCAN-cervix. The MISCAN-Cervix model predicted relatively more excess cancers for women screened with HPV 3-yearly, 5-yearly or 10-yearly due to disruptions, where the second factor seems to outweigh the first. Differences in dwell time for HPV and cervical precancer among the three models contributes to this balance between the two factors (**Appendix**), where the MISCAN-Cervix model has the shortest preclinical dwell time from HPV acquisition to cancer development (20). In addition to the shorter dwelling times, the MISCAN model also assumes that some precancerous lesions are structurally missed over time by cytology-based screening because they are located deeper into the cervical canal. For women with such lesions, missing a screening due to a disruption is less harmful, which reduces the relative difference between primary cytology and primary HPV screening in case of a disruption, and increases the effect of the second factor.

We also found that the relative excess rates of symptomatically-detected cancer projected for the same delay period were higher for women overdue for screening compared with women who attend screening according to guidelines. These relative increases were generally similar regardless of the length of the delay period, as the underlying cases among the guidelines-compliant and overdue women continued to accumulate with the length of delay. However, some delay-length trends were observed (**Figure 2**), which may be due to the fact that the differences in the impact of the delay between guidelines-compliant and under-screeners may become smaller in the case of a longer delay (a longer screening delay increasingly becomes more impactful for guidelines-compliant screeners as well). Delay-length differences are predominantly observed in MISCAN-Cervix, which may be in part due to shorter dwell time assumptions. In contrast, for the Harvard and Policy1-Cervix models that assume longer dwell times, the relative impacts between 1 and 5 year delay is smaller.

Although our analysis was contextualized to the U.S., our results may still be generalizable to other countries where cervical screening was disrupted such as the U.K., Ireland, New Zealand, and the Netherlands where cervical screening was paused for 2-4 months (23-26). We considered combinations of screening modality and screening frequency, some of which will be more applicable to some countries than others. Our overarching findings that disruptions are likely to disproportionately impact those who are already overdue for screening, and that under-screened women are at higher risk than guidelines-compliant screeners affected by a temporary delay are likely generalizable.

Importantly, self-collection of samples at home may provide a tool to reduce screening barriers and facilitate outreach to under-screened people who are also most vulnerable to screening disruptions. In the Netherlands, parts of Sweden, and recently Australia, self-sampling is available to all women, and preliminary findings suggest this has facilitated rapid reintroductions to screening in the Netherlands in the context of COVID-19 (24).

### Limitations and clinical relevance

Despite the strength of consistent results from three established CISNET models, there are several limitations that should be considered in interpreting our results. First, in the absence of detailed information on cervical screening disruptions by screening frequency, we explored a range of stylized scenarios that represent different combinations of screening behavior, disruption periods and screening modalities.

We encountered several challenges as we planned this analysis, which we feel are worth describing as they illustrate some of the limits of modeling in this context. The first is the scope of time over which to consider outcomes. Attempting to assess screening delays over a short period presents problems as short-term outcomes may not be representative of long-term health gains. For instance, the occurrence of screening moments will always be associated with the incidence of cancer due to the volume of screen detected disease. An analysis that attempted to consider changes in the incidence of cancer within a finite period that includes resumption of screening will generate results that are largely artefacts of the resumption of screening within the period of analysis rather than fundamentally reflecting differences due to temporary extensions to the interval. Therefore, we restricted the short-term analysis to the assessment of symptomatically detected cancers during a finite delay period while the long-term analysis considered the net impact on CC risk given women’s lifetime screening participation.

Considering changes in the longer term required us to make assumptions about women’s screening behavior following the COVID-related delays as the impact of the temporary screen delays are contingent on the subsequent screens they receive. Again, we encountered the potential for modelling assumption to influence results. When screening eligibility is limited by an upper age bound, i.e., age 65 years, the impact of a screen delay can depend heavily on what is assumed about a woman’s final screen (Summarized assumptions in **Appendix Tables A1 and A2**). For instance, a 1-year delay to program for a woman following a 5-year interval would imply a final screen occurring at age 61 rather than 65 in the ‘no delay’ scenario. In this instance, the model-projected changes in long-term outcomes potentially reflect a missed final screen moment, rather than isolated only to the extension of 1-year delay (increasing the screening interval temporarily from 5 to 6 years) (22, 27). Changes to future screening patterns maybe an overlooked secondary/long-term consequence of this one-time COVID-19 disruption.

We did not project the impact of temporary disruptions on rarer outcomes such as cancer death as the impact on incidence is an early indicator for mortality burden. We also did not consider differences in underlying risk between various screening behavior groups (i.e., we assumed differential cancer risk was a function of only screening behavior). If women who screen less frequently also face a higher underlying risk of developing cancer, the differences in our projected risks for delays to under-screeners compared with guideline-compliant screeners may be underestimated, providing additional support for a target outreach to under-screened women.

While our models do not explicitly simulate the impacts of specific factors including race and ethnicity, poverty income level, education and insurance status, these characteristics are associated with screening behavior, which we do capture in our simulation models. Furthermore, our projections reflected the burden of cervical cancer assuming an average underlying natural history risk of progression to cervical cancer, and we do not reflect the differential natural history for immunocompromised women. Subsequently, our findings would not be generalizable to certain groups facing greater background risk of developing cancer.

## CONCLUSIONS

Our models predicted that the main driver of lifetime risk of cervical cancer is screening frequency and screening modality, rather than temporary disruptions to screening; however, a disruption to screening does not equally impact women with differential screening histories or screening behaviors. Understanding and reaching under-screened women remains the most critical area of focus, regardless of temporary disruptions.

## Data Availability

Supporting Information contained in the Supplementary Material of Burger et al. (20) provides details on microsimulation model inputs, calibration to epidemiologic data, and calibration approach in line with good modeling practice. Access to the raw results data will be made available upon reasonable request.

## Data availability

Supporting Information contained in the Supplementary Material of Burger et al. (20) provides details on microsimulation model inputs, calibration to epidemiologic data, and calibration approach in line with good modeling practice. This study involved modelling rather than direct analysis of primary datasets. The current manuscript is a computational study, so no data have been generated for this manuscript. The Cancer Intervention and Surveillance Modeling Network (CISNET) (https://cisnet.cancer.gov/) Cervix model codes have been developed over decades, are proprietary property, and cannot be provided by the authors at this time; however, CISNET-Cervix, under our CISNET ‘Model Accessibility’ interest group is working to provide transparent and reproducible modeling code for forthcoming projects (“C4”). Access to current code is possible only through supervised training at each modeling group cite.

## APPENDIX

### 1. Analytic screening assumptions

There were differences in how each modeling group applied screening. In order to isolate health outcomes to women disrupted in 2020, the Harvard and MISCAN-Cervix models assumed primary screening followed fixed intervals (so-called “age-based” screening) irrespective of possible follow-up testing which would normally affect the age at which a woman would next screen. In contrast, the Policy1-Cervix model allowed for dynamic primary screening based on time since last primary or follow-up screen, but isolated their model outcomes to the women who were delayed in 2020, i.e., did not have previous positive results. Subsequently, the Policy-1-Cervix model reflects a marginally lower-risk cohort of women; however, while the absolute lifetime risk may be lower compared with the other two models, absolute changes in risk compared with their counterfactual of “no delay” will isolate the impact of delays.

In line with US guideline recommendations, all models assumed screening ended no later than age 65 years (inclusive). Scenarios involving delays to screening shifted the timing of future screening in all groups other than annual screeners, and or some screening frequency and delay combinations, the delays to screening reduced the number of lifetime screens and changed the implied age of last screen (due to fixed screening intervals post-delay) (**Appendix Tables A1 and A2**). For example, for 10-yearly screeners in the 1975 birth cohort (age 45 in 2020), a 1-year delay meant their last screening test occurred at age 56, 9 years earlier than in the ‘no delay’ scenario. In these same 10-yearly screeners, a 2- and 5-year delay shifted their age at last screen to age 57 and 60 years, respectively. Some of the added risk of the longer delay compared with a 1-year delay may be mitigated by the later screening end age in the longer delay scenarios (that end screening at age 57 or 60, compared to age 56). Model-based analyses require making analytic assumptions about imperfect screening behavior and guidelines.

**Table A1.** Screening end age (lifetime number of screens) by birth cohort, screening frequency and delay duration.

|  |  | Annual | 3-yearly* | 5-yearly** | 10-yearly |
| --- | --- | --- | --- | --- | --- |
| 1965<br>(age 55 in<br>2020) | No delay | 65 (45) | 64 (15) | 65 (9) | 65 (5) |
|  | 1-year delay | 65 (44) | 65 (15) | 61 (8) | 56 (4) |
|  | 2-year delay | 65 (43) | 63 (14) | 62 (8) | 57 (4) |
|  | 5-year delay | 65 (40) | 63 (13) | 65 (8) | 60 (4) |
| 1975<br>(age 45 in<br>2020) | No delay | 65 (45) | 63 (15) | 65 (9) | 65 (5) |
|  | 1-year delay | 65 (44) | 64 (15) | 61 (8) | 56 (4) |
|  | 2-year delay | 65 (43) | 65 (15) | 62 (8) | 57 (4) |
|  | 5-year delay | 65 (40) | 65 (14) | 65 (8) | 60 (4) |
| 1985<br>(age 35 in<br>2020) | No delay | 65 (45) | 65 (15) | 65 (8) | 65 (5) |
|  | 1-year delay | 65 (44) | 63 (14) | 61 (8) | 56 (4) |
|  | 2-year delay | 65 (43) | 64 (14) | 62 (8) | 57 (4) |
|  | 5-year delay | 65 (40) | 64 (13) | 65 (8) | 60 (4) |

\*Guidelines-compliant screener with primary cytology-based screening;
\*\*Guidelines-compliant screener with primary human papillomavirus (HPV) testing for women aged 30+ years.

**Table A2.**
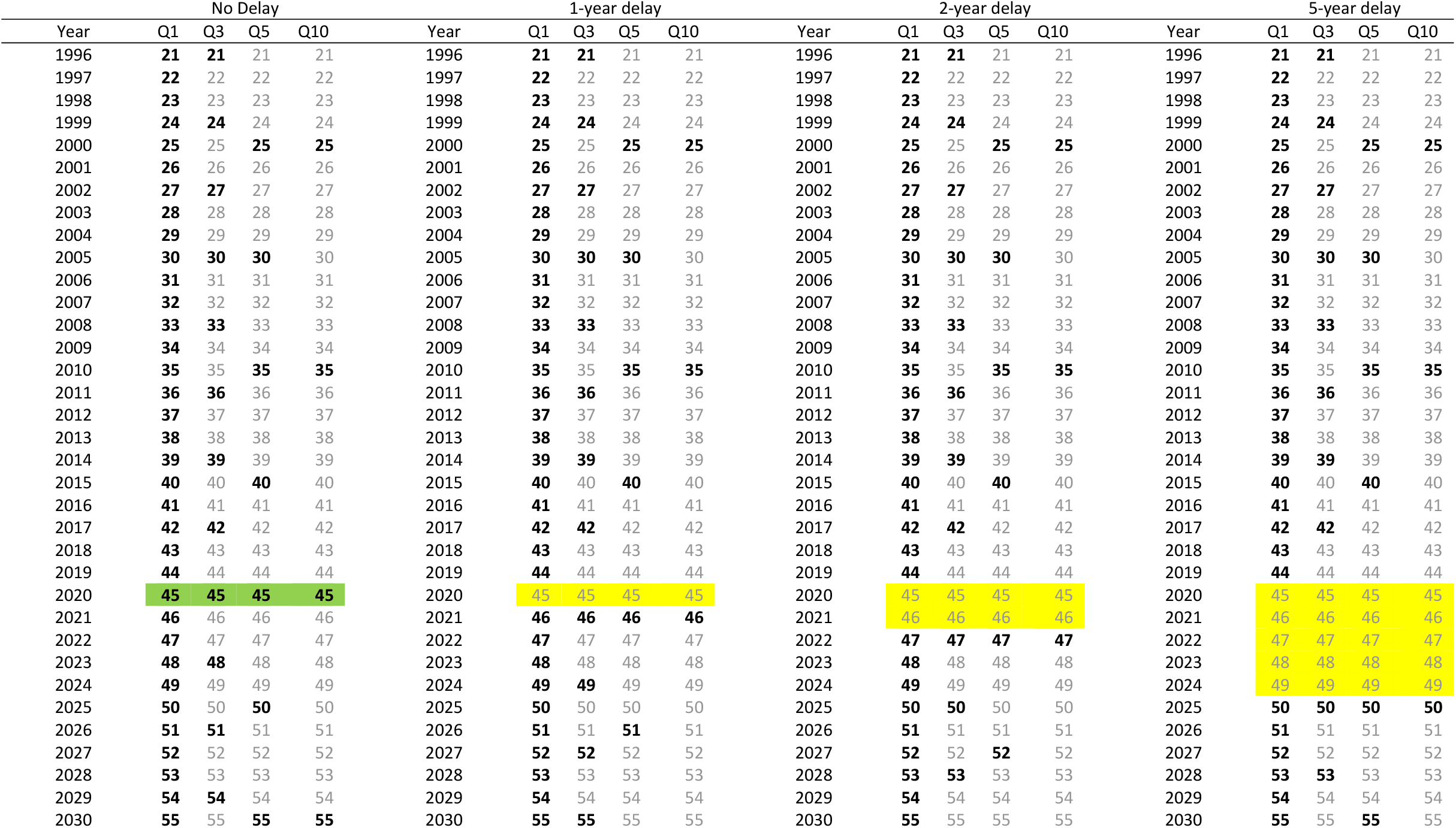
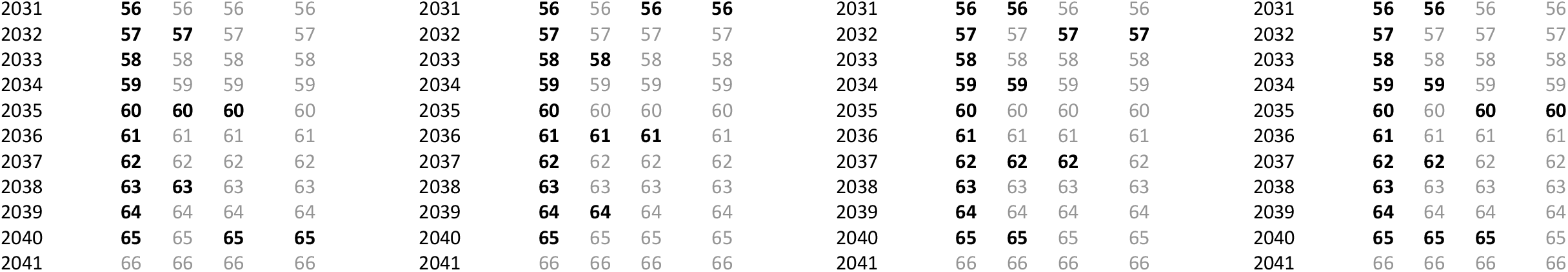
Example age at screen for the 1975 birth cohort without (highlighted in green) and with (highlighted in yellow) COVID-19-related delays, by screening frequency. Numbers under each delay are ages, bolded numbers are ages at which screening takes place, green highlight reflects no delay, and yellow highlights reflects a delay.

**Figure A1.**
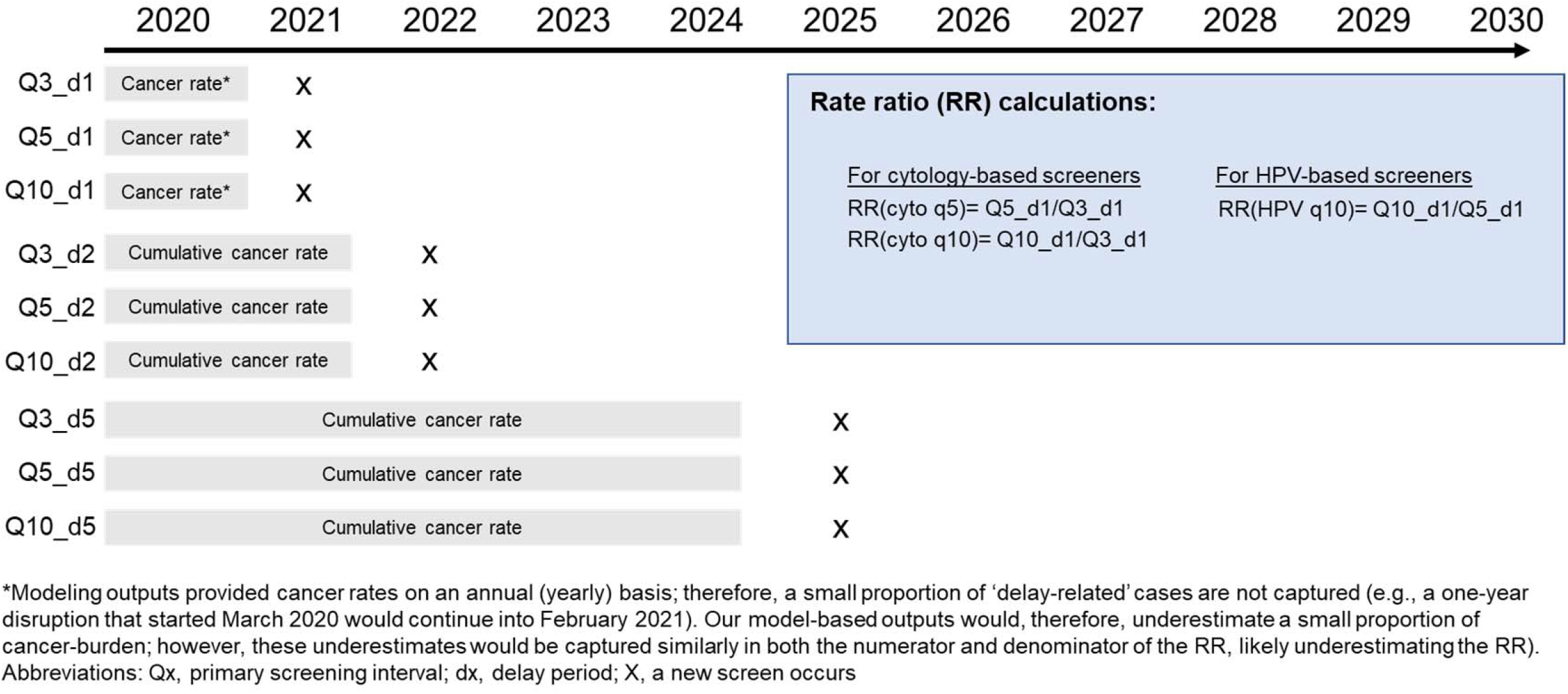
Schematic of short-term cancer burden calculations*

### 2. Additional results

**Table A3.** Relative rate ratios and accumulated incidence rates per 100,000 women for each screening frequency and delay scenario

| Screening frequency | 1-yr delay | 2-yr delay | 5-year delay |
| --- | --- | --- | --- |
| *Policy1-Cervix* |  |  |  |
| 5-yearly screener (cytology) | 1.40 = 4.94 / 3.58 | 1.44 = 10.05 / 7.00 | 1.57 = 30.03 / 19.10 |
| 10-yearly screener (cytology) | 3.10 = 11.06 / 3.58 | 3.35 = 23.45 / 7.00 | 3.34 = 63.72 / 19.10 |
| 10-yearly screener (HPV) | 2.26 = 7.36 / 3.26 | 2.48 = 15.31 / 6.18 | 2.54 = 42.41 / 16.67 |
| *MISCAN-Cervix* |  |  |  |
| 5-yearly screener (cytology) | 2.78 = 3.00 / 1.08 | 2.28 = 6.41 / 2.81 | 1.99 = 22.09 / 11.08 |
| 10-yearly screener (cytology) | 6.97 = 7.54 / 1.08 | 5.73 = 16.11 / 2.81 | 4.15 = 45.97 / 11.08 |
| 10-yearly screener (HPV) | 3.91 = 6.08 / 1.55 | 3.67 = 13.08 / 3.56 | 2.97 = 38.84 / 13.09 |
| *Harvard* |  |  |  |
| 5-yearly screener (cytology) | 1.8 = 9.64 / 5.37 | 1.74 = 17.69 / 10.15 | 1.64 = 45.97 / 27.96 |
| 10-yearly screener (cytology) | 3.8 = 20.39 / 5.37 | 3.64 = 36.94 / 10.15 | 3.19 = 89.29 / 27.96 |
| 10-yearly screener (HPV) | 3.61 = 10.74 / 2.98 | 3.58 = 19.51 / 5.45 | 3.69 = 46.83 / 12.68 |

Relative rate ratio is calculated as the accumulated incidence rate per 100,000 women during a delay period for a give screening history divided by the accumulated incidence rate per 100,000 women during the same delay period among guidelines-compliant screeners. Incidence rates are the average across the three birth cohorts. 3-yearly cytology screening is considered guidelines-compliant screening; 5-yearly HPV screening is considered guidelines compliant.

**Table A4.** Percentage reduction in average (across the 1965, 1975, and 1985 birth cohorts) lifetime risk of cancer compared with no screening

|  |  | Screening frequency |  |  |  |
| --- | --- | --- | --- | --- | --- |
|  |  | Annual | 3-yearly | 5-yearly | 10-yearly |
| **Primary cytology-based screening** |  |  |  |  |  |
| *Harvard* |  |  |  |  |  |
|  | No delay | 88.4 % | 79.9 % | 71.4 % | 56.3 % |
|  | 1-year delay | 88.5 % | 80.0 % | 70.9 % | 55.3 % |
|  | 2-year delay | 88.4 % | 79.5 % | 70.5 % | 54.9 % |
|  | 5-year delay | 87.9 % | 78.4 % | 69.3 % | 53.4 % |
| *MISCAN-Cervix* |  |  |  |  |  |
|  | No delay | 85.1 % | 72.1 % | 62.1 % | 48.4 % |
|  | 1-year delay | 85.0 % | 71.7 % | 60.2 % | 44.7 % |
|  | 2-year delay | 84.7 % | 71.2 % | 59.9 % | 44.9 % |
|  | 5-year delay | 83.1 % | 69.1 % | 58.8 % | 43.9 % |
| *Policy1-Cervix* |  |  |  |  |  |
|  | No delay | 87.7 % | 86.5 % | 84.5 % | 75.9 % |
|  | 1-year delay | 87.8 % | 86.3 % | 82.3 % | 71.9 % |
|  | 2-year delay | 87.7 % | 85.8 % | 82.5 % | 71.9 % |
|  | 5-year delay | 86.8 % | 85.1 % | 82.5 % | 71.3 % |
| **Primary HPV-based screening** |  |  |  |  |  |
| *Harvard* |  |  |  |  |  |
|  | No delay | 92.6 % | 89.3 % | 86.7 % | 77.8 % |
|  | 1-year delay | 92.6 % | 89.4 % | 86.6 % | 77.1 % |
|  | 2-year delay | 92.6 % | 89.4 % | 86.6 % | 76.7 % |
|  | 5-year delay | 92.6 % | 89.2 % | 86.4 % | 75.0 % |
| *MISCAN-Cervix* |  |  |  |  |  |
|  | No delay | 94.0 % | 84.2 % | 74.8 % | 58.0 % |
|  | 1-year delay | 93.9 % | 83.8 % | 72.3 % | 53.5 % |
|  | 2-year delay | 93.6 % | 83.2 % | 72.3 % | 53.5 % |
|  | 5-year delay | 92.3 % | 81.0 % | 70.8 % | 51.8 % |
| *Policy1-Cervix* |  |  |  |  |  |
|  | No delay | 90.9 % | 90.4 % | 90.1 % | 86.6 % |
|  | 1-year delay | 90.9 % | 90.4 % | 88.7 % | 82.7 % |
|  | 2-year delay | 91.0 % | 90.1 % | 89.0 % | 82.8 % |
|  | 5-year delay | 91.0 % | 90.0 % | 89.2 % | 82.6 % |

